# COVID-19 ICU and mechanical ventilation patient characteristics and outcomes - A systematic review and meta-analysis

**DOI:** 10.1101/2020.08.16.20035691

**Authors:** Raymond Chang, Khaled Mossad Elhusseiny, Yu-Chang Yeh, Wei-Zen Sun

## Abstract

**Background:** Insight into COVID-19 intensive care unit (ICU) patient characteristics, rates and risks of invasive mechanical ventilation (IMV) and associated outcomes as well as any regional discrepancies is critical in this pandemic for individual case management and overall resource planning.

**Methods and Findings:** Electronic searches were performed for reports through May 1 2020 and reports on COVID-19 ICU admissions and outcomes were included using predefined search terms. Relevant data was subsequently extracted and pooled using fixed or random effects meta-analysis depending on heterogeneity. Study quality was assessed by the NIH tool and heterogeneity was assessed by I^2^ and Q tests. Baseline patient characteristics, ICU and IMV outcomes were pooled and meta-analyzed. Pooled odds ratios (pOR) were calculated for clinical features against ICU, IMV mortality. Subgroup analysis was carried out based on patient regions. A total of twenty-eight studies comprising 12,437 COVID-19 ICU admissions from seven countries were meta-analyzed. Pooled ICU admission rate was 21% [95% CI 0.12-0.34] and 69% of cases needed IMV [95% CI 0.61-0.75]. ICU and IMV mortality were 28.3% [95% CI 0.25-0.32], 43% [95% CI 0.29-0.58] and ICU, IMV duration was 7.78 [95% CI 6.99-8.63] and 10.12 [95% CI 7.08-13.16] days respectively. Besides confirming the significance of comorbidities and clinical findings of COVID-19 previously reported, we found the major correlates with ICU mortality were IMV [pOR 16.46, 95% CI 4.37-61.96], acute kidney injury (AKI) [pOR 12.47, 95% CI 1.52-102.7], and acute respiratory distress syndrome (ARDS) [pOR 6.52, 95% CI 2.66-16.01]. Subgroup analyses confirm significant regional discrepancies in outcomes.

**Conclusions:** This is the most comprehensive systematic review and meta-analysis of COVID-19 ICU and IMV cases and associated outcomes to date and the only analysis to implicate IMV’s associtaion with COVID-19 ICU mortality. The significant association of AKI, ARDS and IMV with mortality has implications for ICU resource planning for AKI and ARDS as well as research into optimal ventilation strategies for patients. Regional differences in outcome implies a need to develop region specific protocols for ventilatory support as well as overall treatment.

## INTRODUCTION

Since its inception, the Coronavirus 2019 (COVID-19) pandemic has placed unprecedented strain on healthcare globally with intensive care unit (ICU) care and ventilator availability being focal points of health resource allocation and planning. Understanding COVID-19 ICU and associated invasive mechanical ventilation (IMV) patient characteristics and outcomes as well as appreciating their regional variations is thus critically important for patient management as well as resource planning. Despite an increasing number of studies relating to various aspects of severe COVID-19 and its ICU management; discrepancies in size, methodologies and research focus as well as regional differences in these studies calls for a comprehensive systematic review and analysis of reports to date.

## METHODS

### Search strategy

Our review was conducted according to PRISMA guidelines [1] (S1 Table). We searched Pubmed, Scopus, Embase, preprint servers bioRvix and medRvix and the Intensive Care National Audit and Research Center (ICNARC) website for publications through May 1^st^ 2020. A hand search through referenced published studies was additionally performed. Search terms used were (“COVID-19” OR “SARS-Cov-2”) AND (“ICU” or “ventilator” OR “mechanical ventilation” OR “critical care” OR “intensive care” OR “hospitalized” OR “outcomes”). The study protocol was registered with the International Prospective Register of Systematic Reviews (PROSPERO CRD42020182482).

### Selection criteria

Studies reporting COVID-19 cases and their ICU admissions and IMV outcomes were included. Case reports, abstracts, or studies without outcomes were excluded. Two authors (RC and KME) independently screened titles and abstracts of retrieved studies and reviewed full-texts for further inclusion. Studies with larger outcome samples from same hospitals were analyzed to avoid patient overlaps; however, studies with overlapped patients but distinct outcome measures were meta-analyzed for those outcomes.

### Data extraction

Data extracted independently by two authors (RC and KME) into Excel (Microsoft) included study characteristics and patient clinical characteristics, and outcomes.

### Quality assessment

Two authors (RC and KME) independently assessed the risk of bias of included studies utilizing the NIH quality assessment tool for observational cohorts [2]. The overall assessment was good, fair, or poor. Disagreements were resolved by discussion between all authors.

### Data analysis

Meta-analysis was conducted when there were two or more studies for the same outcome using Comprehensive Meta-analysis (CMA) software version 3. The pooled event rate (pER) was used to meta-analyze the prevalences of patient characteristics, rates of ICU admissions and IMV, and ICU and IMV mortalities. Pooled odds ratio (pOR) was used to assess the association between clinical features and outcomes. Pooled mean or weighted mean differences with 95% confidence intervals (95% CI) were used in meta-analysis of continuous outcomes. We used the fixed-effect model unless there was evidence of heterogeneity, in which case the random-effects model was used. Heterogeneity was determined using I^2^ and Q test [3.4] and was considered significant if the P-value of the Q test is <0.1 and/or I^2^ >50%. The corresponding 95% CI of the pooled effect size was calculated. Subgroup analysis were done on patient region, overall quality, and publication type (pre-print vs. peer-reviewed) to investigate the heterogeneity amongst studies. Publication bias was assessed using Egger’s regression test and funnel plots for outcomes with five or more studies, and was considered significant if Egger’s regression P-value is <0.1 [5,6]. When a mean value of a continuous outcome was missing, methods by Wan [7] were utilized to impute the needed value for the meta-analysis. Since some studies [8,9,10-17,18-23] included patients still hospitalized at endpoint (n=4,697, 37.8%), we considered two mortality scenarios methodologically: (1) best case scenario by pooling patients with known outcome and (2) worst case scenario by pooling those still hospitalized with non-survivors.

## RESULTS

### Search results

We retrieved 7846 studies and removed 5134 duplicates and remaining articles were screened to yield 49 studies for full-text review (Figure 1). Twenty-eight studies were finally included and 16 were peer-reviewed, 11 were preprints, while one was an online report [24].

**Fig 1.**
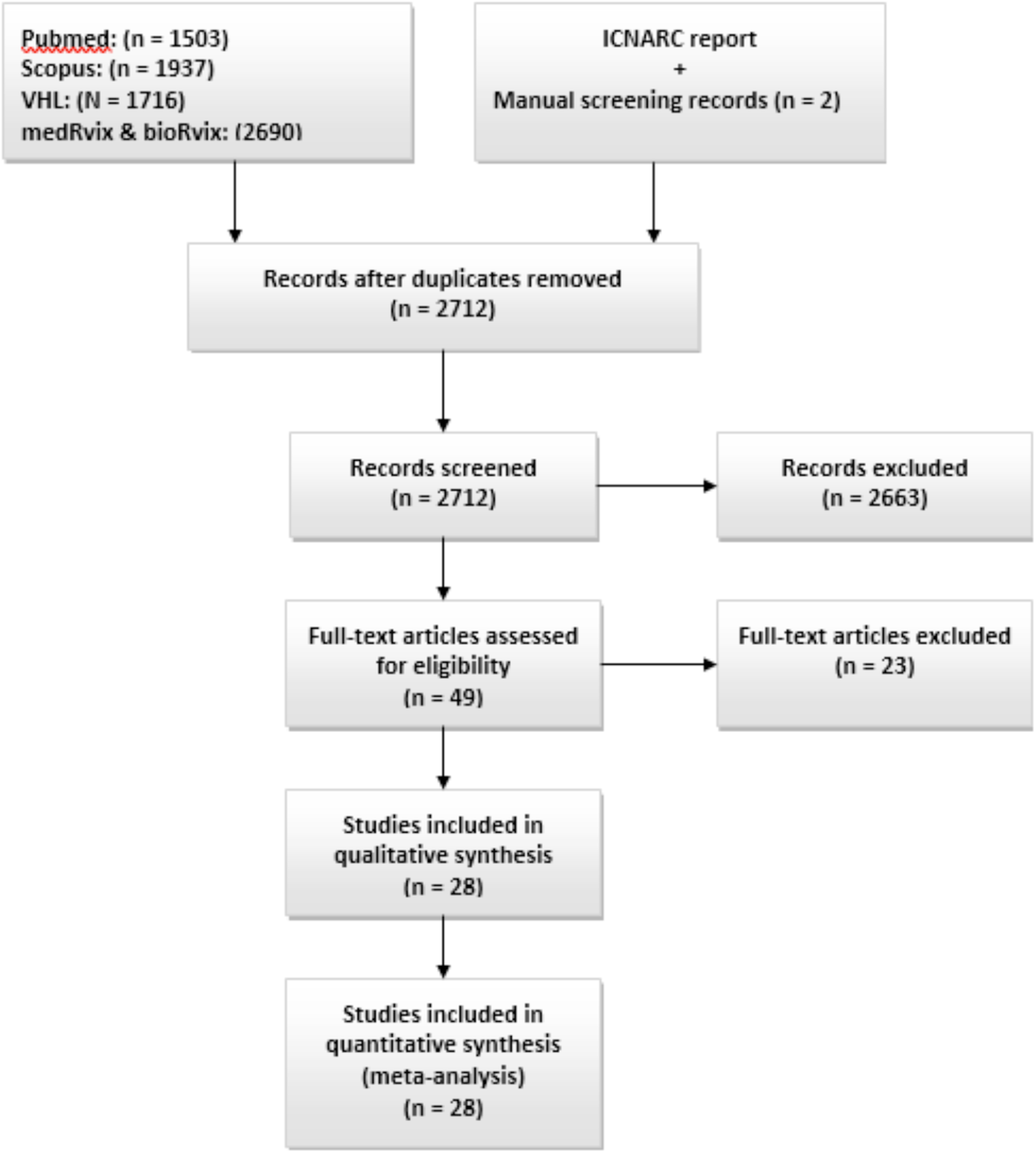
PRISMA flow diagram. PRISMA diagram depicting study selection process

### Characteristics of studies and patients (Table 1)

All included studies were observational and comprised 12,437 ICU patients admitted between December 2019 to May 1 2020. Of these, 6,875 patients were on IMV. Nine studies were from the USA [8-11,17,18,23,25,26], 13 from China [12,14,16,20-22,27-33], two from the UK [24,32], and one each from Italy [15], Spain [13], France [19], and Mexico [35].

### Risk of bias assessment (Table 1)

Fifteen studies were good and 13 were fair as assessed by the NIH tool. Follow-up duration was insufficient or unreported in seven [8,10,12,14,26,29,35] among which three had fair assessments. Of note, 14 studies [8,9,12-15,17-19,21,22,24,25,34] had over 20% of patients with unknown outcome at endpoint of which 8 had fair assessment.

**Table 1.**
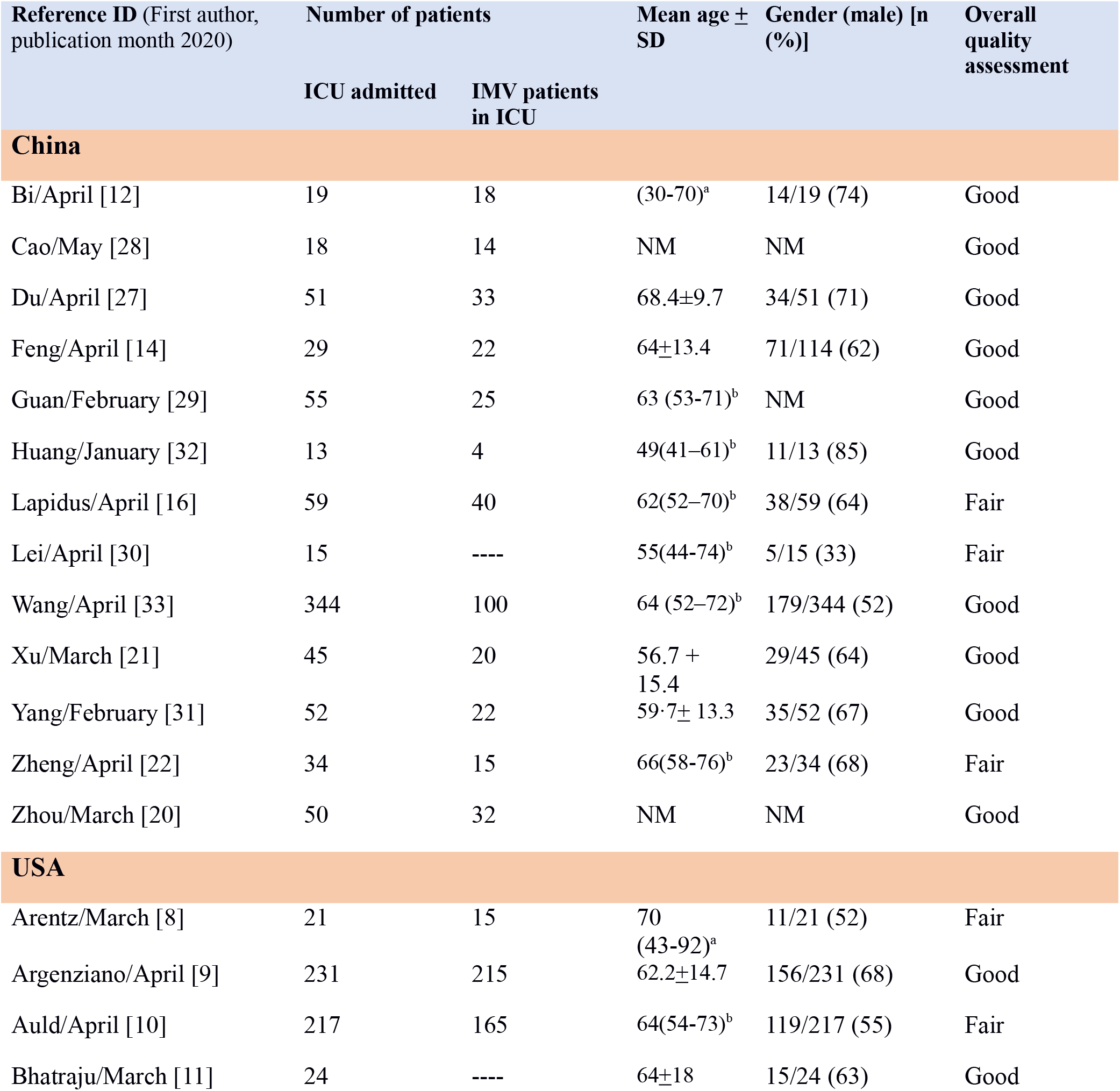
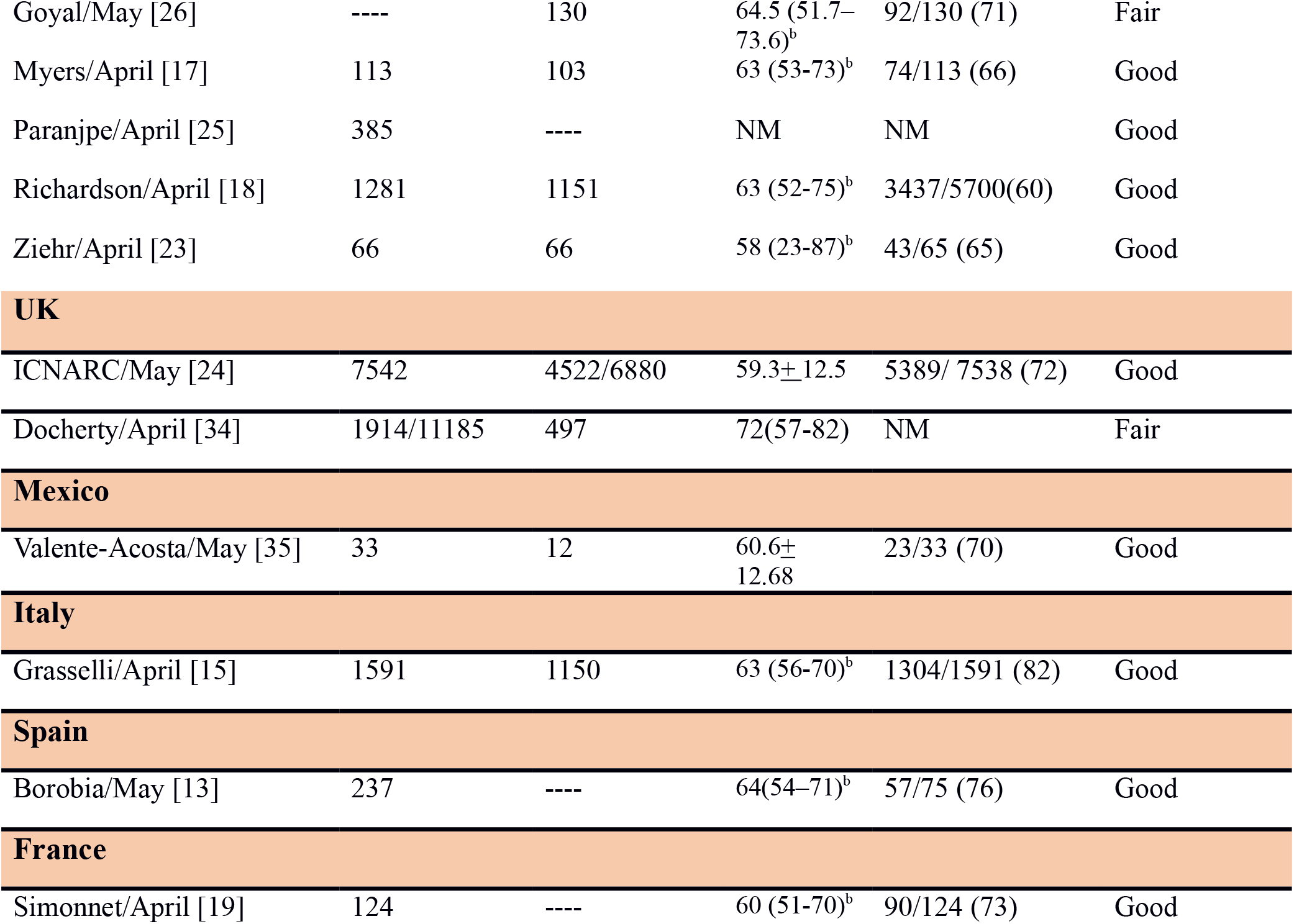
COVID-19 ICU study regions, ICU admitted and IMV patient number and demographics, and study quality assessement. Studies based on reporting region and identified by first author and 2020 publication month, the number of ICU admitted and IMV patients in ICU and their age and sex as reported per study; and overall quality assessement of the studies by the NIH quality assessment tool [2]. Abbreviations: NM= Not mentioned, a = Data range, b =Data are reported as median (IQR).

| Reference ID (First author, publication month 2020) | Number of patients | | Mean age ± SD | Gender (male) [n (%)] | Overall quality assessment |
| --- | --- | --- | --- | --- | --- |
|  | ICU admitted | IMV patients in ICU |  |  |  |
| China |  |  |  |  |  |
| Bi/April [12] | 19 | 18 | (30-70)<sup>a</sup> | 14/19 (74) | Good |
| Cao/May [28] | 18 | 14 | NM | NM | Good |
| Du/April [27] | 51 | 33 | 68.4±9.7 | 34/51 (71) | Good |
| Feng/April [14] | 29 | 22 | 64±13.4 | 71/114 (62) | Good |
| Guan/February [29] | 55 | 25 | 63 (53-71)<sup>b</sup> | NM | Good |
| Huang/January [32] | 13 | 4 | 49(41-61)<sup>b</sup> | 11/13 (85) | Good |
| Lapidus/April [16] | 59 | 40 | 62(52-70)<sup>b</sup> | 38/59 (64) | Fair |
| Lei/April [30] | 15 | ---- | 55(44-74)<sup>b</sup> | 5/15 (33) | Fair |
| Wang/April [33] | 344 | 100 | 64 (52-72)<sup>b</sup> | 179/344 (52) | Good |
| Xu/March [21] | 45 | 20 | 56.7 + 15.4 | 29/45 (64) | Good |
| Yang/February [31] | 52 | 22 | 59.7± 13.3 | 35/52 (67) | Good |
| Zheng/April [22] | 34 | 15 | 66(58-76)<sup>b</sup> | 23/34 (68) | Fair |
| Zhou/March [20] | 50 | 32 | NM | NM | Good |
| USA |  |  |  |  |  |
| Arentz/March [8] | 21 | 15 | 70 (43-92)<sup>a</sup> | 11/21 (52) | Fair |
| Argenziano/April [9] | 231 | 215 | 62.2±14.7 | 156/231 (68) | Good |
| Auld/April [10] | 217 | 165 | 64(54-73)<sup>b</sup> | 119/217 (55) | Fair |
| Bhatraju/March [11] | 24 | ---- | 64±18 | 15/24 (63) | Good |
| Goyal/May [26] | ---- | 130 | 64.5 (51.7–73.6)<sup>b</sup> | 92/130 (71) | Fair |
| Myers/April [17] | 113 | 103 | 63 (53-73)<sup>b</sup> | 74/113 (66) | Good |
| Paranjpe/April [25] | 385 | ---- | NM | NM | Good |
| Richardson/April [18] | 1281 | 1151 | 63 (52-75)<sup>b</sup> | 3437/5700(60) | Good |
| Ziehr/April [23] | 66 | 66 | 58 (23-87)<sup>b</sup> | 43/65 (65) | Good |
| UK |  |  |  |  |  |
| ICNARC/May [24] | 7542 | 4522/6880 | 59.3±12.5 | 5389/ 7538 (72) | Good |
| Docherty/April [34] | 1914/11185 | 497 | 72(57-82) | NM | Fair |
| Mexico |  |  |  |  |  |
| Valente-Acosta/May [35] | 33 | 12 | 60.6±12.68 | 23/33 (70) | Good |
| Italy |  |  |  |  |  |
| Grasselli/April [15] | 1591 | 1150 | 63 (56-70)<sup>b</sup> | 1304/1591 (82) | Good |
| Spain |  |  |  |  |  |
| Borobia/May [13] | 237 | ---- | 64(54–71)<sup>b</sup> | 57/75 (76) | Good |

**Table 1. COVID-19 ICU study regions, ICU admitted and IMV patient number and demographics , and study quality assessement.**
|  |  |  |  |  |  |
| --- | --- | --- | --- | --- | --- |
| Simonnet/April [19] | 124 | ---- | 60 (51-70)<sup>b</sup> | 90/124 (73) | Good |

## Quantitative analysis

### ICU admissions, duration and outcome

Pooled ICU admission rate among 17,639 hospitalized COVID-19 patients meta-analyzed from eight studies [9,12,14,18,25,27,29,34] was 21% (95% CI 0.12-0.34) (Fig 2a) and pooled ICU mortality rate of 12,437 patients from 20 studies [8-19,21-25,31,33,35] was 28.3% (95% CI 0.27-0.36) (Fig 2b), while the worst case scenario mortality is 60% (95% CI 0.49-0.69) considering 4,697 patients with unknown outcomes. ICU length of stay (LoS) from five studies [8,10,11,15,24] had a pooled mean duration of 7.78 (7.05-8.51) days. Substantial heterogeneity was observed with ICU outcomes (p-value <0.1, I^2^ > 60%) and not explained by subgroup analysis. Eggers’ test revealed no publication bias (Figure 3).

**Fig 2.**
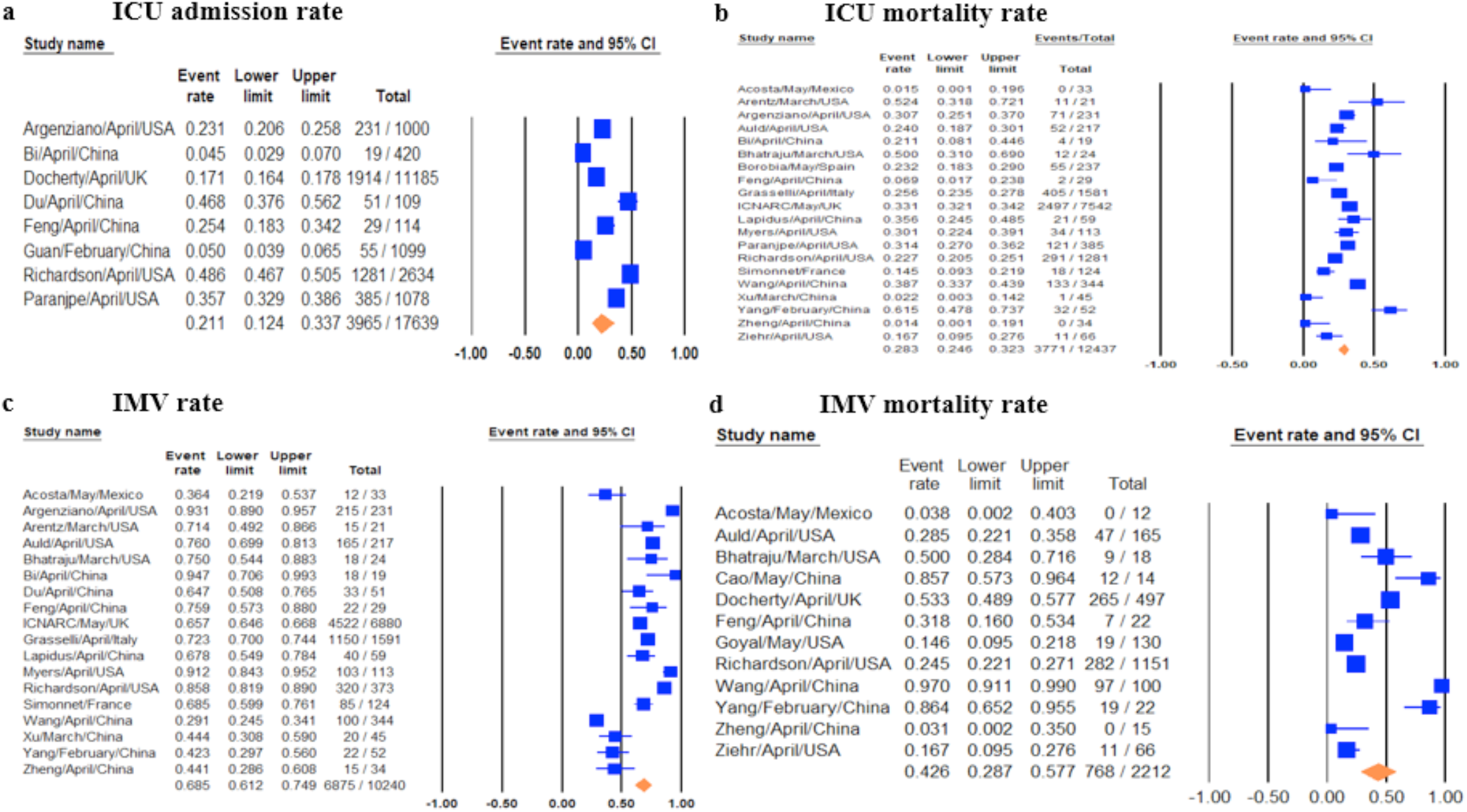
Forest plots of relevant included studies showing pooled event rates of ICU and IMV outcomes. Forest plots of studies for 2a) ICU admission rates, 2b) ICU mortality rates, 2c) IMV rates, 2d) IMV mortality rates.

**Fig 3.**
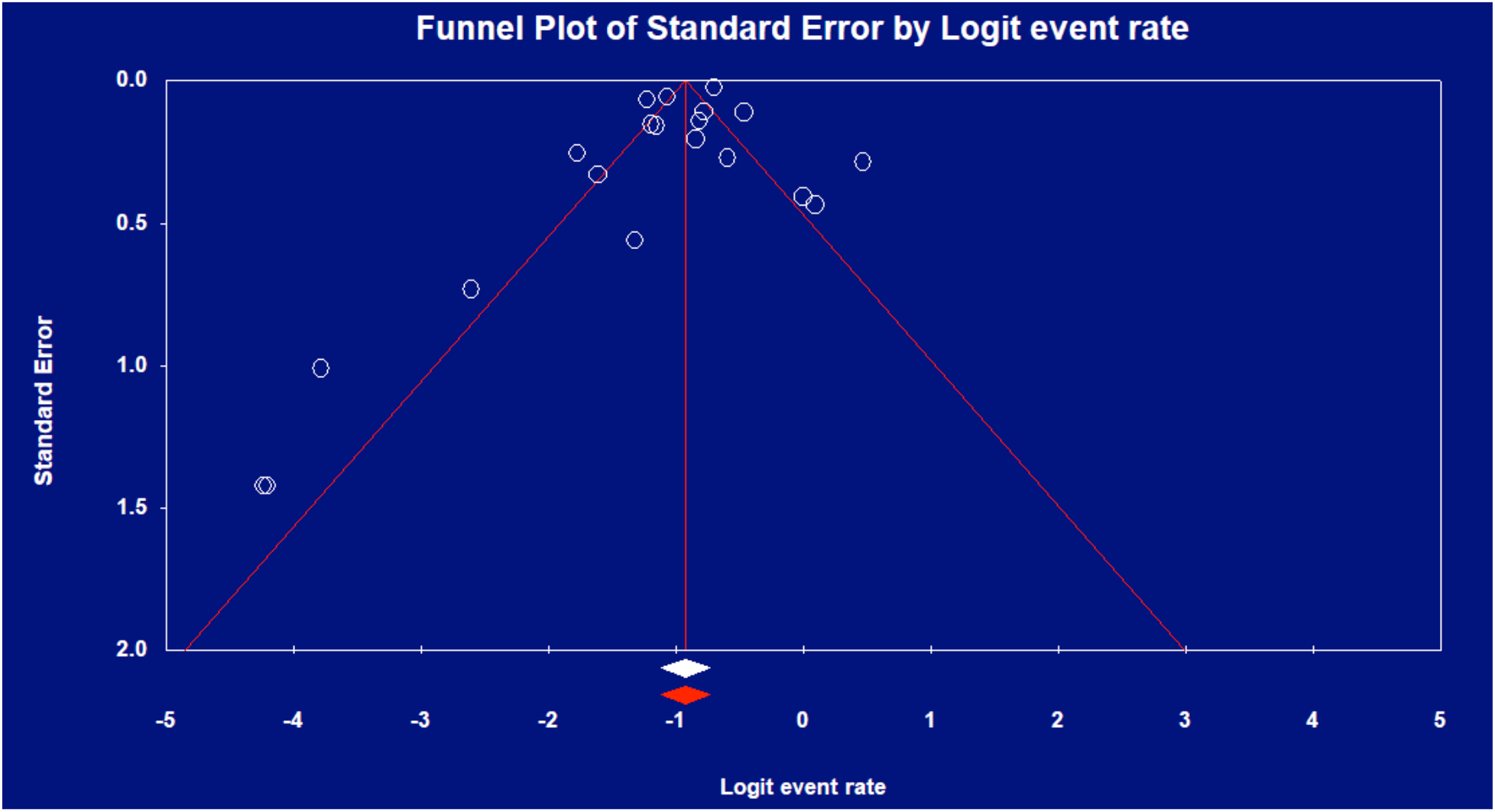
Funnel plot of ICU mortality rate.

### IMV prevalence, duration and outcome

Pooled IMV prevalence and mortality was respectively 69% (95% CI 0.61-0.75) from 18 studies [8,9-12,15-19,21,22,24,27,31,33,35] with 10,240 cases (Figs 2c) and 43% (95% CI 0.29-0.58) from 12 studies [10,11,14,18,22,23,26,28,31,33-35] with 2,212 cases based on a best case scenario, with worst case scenario mortality of 74% (95% CI 0.54-0.87) (Fig. 2d). IMV duration was pooled from four studies [9,15-17] with a mean duration of 10.12 days (95% CI 7.08-13.16). There was significant heterogeneity among IMV mortalities (p-value < 0.001, I^2^ >90%), which was not explained by subgroup analysis, and Egger’s test revealed no publication bias (Fig 4).

**Fig 4.**
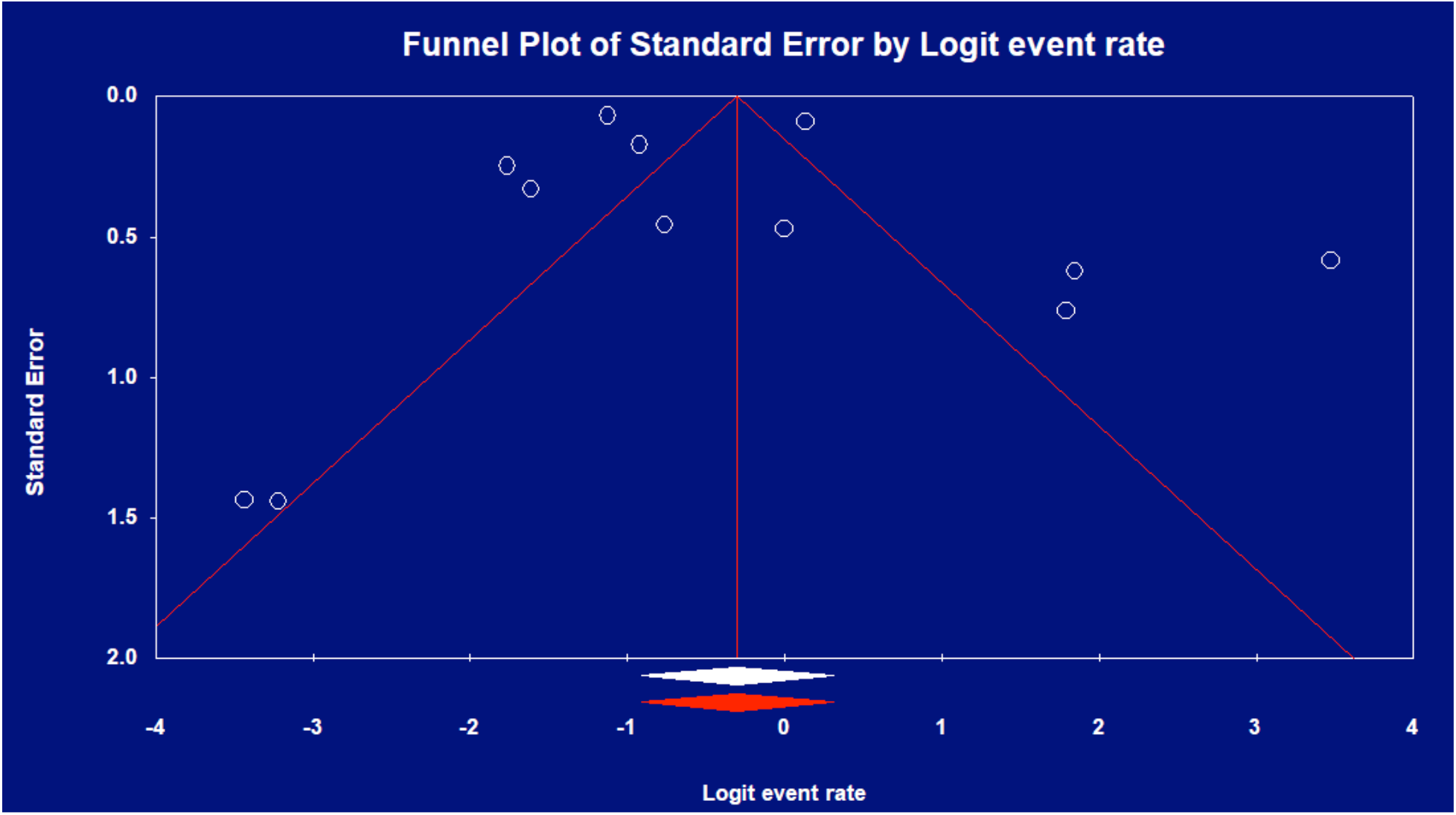
Funnel plot of IMV mortality rate.

#### Prevalence of comorbidities and clinical features

The prevalence of comorbidities as well as presenting clinical features are listed in Table 2 with hypertension (HTN) 51%, obesity (BMI>30kg/m^2^) 35%, diabetes (DM) 30% and fever 81%, cough 76%, dyspnea 75% the 3 most prevalent comorbidities and symptoms respectively. The pER’s of select reported test findings were: bilateral infiltrates on chest radiography 83%, lymphopenia 78%, elevated alanine (ALT), aspartate aminotransferases (AST) 71%, 66.3% respectively, elevated troponin 22%. The pER for concurrent acute respiratory distress syndrome (ARDS) and acute kidney injury (AKI) were 84% and 32% respectively.

**Table 2.** Pooled prevalence of comorbidities and clinical features among ICU-admitted COVID-19 patients. Pooled prevalence of COVID-19 ICU admitted patients’ clinical characteristics including comorbidities, symptoms, signs, laboratory and radiographic findings, as well as complications of ARDS and AKI. Abbreviations: ALT, alanine aminotransferase; AST, aspartate aminotransferase; AKI, acute kidney injury; ARDS, acute respiratory distress syndrome; COPD, chronic obstructive pulmonary disease; CVD, cardiovascular disease.

| Clinical characteristics | Number of studies pooled | Heterogeneity |  | Prevalence (95%CI) |
| --- | --- | --- | --- | --- |
|  |  | P value | I<sup>2</sup> |  |
| Comorbidities |  |  |  |  |
| Hypertension | 15 | <0.01 | 78.81 | 0.51 (0.46 - 0.56) |
| Obesity | 5 | <0.01 | 94.85 | 0.35 (0.23 - 0.49) |
| Diabetes mellitus | 17 | <0.01 | 89.74 | 0.29 (0.23 - 0.37) |
| Respiratory viral co-infection | 3 | <0.01 | 94.46 | 0.21 (0.004 - 0.94) |
| CHF | 5 | <0.01 | 88.36 | 0.16 (0.10 - 0.25) |
| Smoking | 6 | <0.01 | 90.14 | 0.15 (0.07 - 0.31) |
| CVD | 3 | 0.4 | 00.00 | 0.13 (0.104 - 0.17) |
| Obstructive sleep apnea | 3 | <0.01 | 90.43 | 0.13 (0.03 - 43) |
| Asthma | 5 | 0.4 | 00.00 | 0.103 (0.08 - 0.13) |
| Chronic Kidney disease | 15 | <0.01 | 96.97 | 0.09 (0.04 - 0.18) |
| COPD | 12 | <0.01 | 78.08 | 0.09 (0.06 - 0.13) |
| Malignancy | 13 | <0.01 | 93.33 | 0.07 (0.04 - 0.12) |
| Immuno-suppressive therapy | 5 | 0.04 | 73.87 | 0.06 (0.03 - 0.12) |
| History of organ transplantation | 2 | 0.4 | 00.00 | 0.05 (0.03 - 0.09) |
| Liver disease | 11 | <0.01 | 90.99 | 0.03 (0.01 - 0.07) |
| HIV | 3 | 0.7 | 00.00 | 0.03 (0.01 - 0.05) |
| **Complications** |  |  |  |  |
| ARDS | 6 | <0.01 | 96.13 | 0.84 (0.59 – 0.95) |
| AKI | 6 | <0.01 | 96.49 | 0.32 (0.13 – 0.58) |
| **Symptoms and signs** |  |  |  |  |
| Fever | 11 | <0.01 | 92.95 | 0.81 (0.67 – 0.89) |
| Cough | 12 | <0.01 | 89.08 | 0.76 (0.56 – 0.77) |
| Shortness of breath | 10 | <0.01 | 82.21 | 0.75 (0.66 – 0.81) |
| Malaise | 3 | <0.01 | 89.39 | 0.53 (0.25 – 0.79) |
| Fatigue | 5 | <0.01 | 82.76 | 0.46 (0.32 – 0.61) |
| Myalgia | 8 | <0.01 | 83.26 | 0.23 (0.14 – 0.36) |
| Diarrhea | 9 | 0.03 | 52.71 | 0.23 (0.17 – 0.29) |
| Sore throat | 2 | 0.1 | 62.68 | 0.12 (0.05 – 0.27) |
| Nausea and vomiting | 5 | 0.06 | 54.94 | 0.11 (0.07 – 0.19) |
| Headache | 9 | <0.01 | 79.45 | 0.11 (0.05 – 0.22) |
| Rhinorrhea | 4 | 0.5 | 0 | 0.09 (0.06 – 0.13) |
| **Investigational findings** |  |  |  |  |
| Bilateral infiltrates on chest radiography | 6 | <0.01 | 96.73 | 0.83 (0.53 - 0.96) |
| Lymphocytes, <1000/μL | 6 | 0.09 | 47.49 | 0.78(0.72 - 0.84) |
| AST, >40/L | 4 | 0.03 | 65.55 | 0.58 (0.35 – 0.75) |
| ALT, >40/L | 3 | <0.01 | 87.26 | 0.47 (0.14 – 0.83) |
| Troponin, >99th percentile | 2 | 0.3 | 22.04 | 0.22 (0.11 – 0.39) |

#### Association of patient characteristics with ICU and IMV mortality

We analyzed associated COVID-19 ICU patient characteristics and initial laboratory findings for potential association with ICU (Table 3 and Table 4) and IMV survival. Demographically, age>60 years (pOR 3.7, 95% CI 2.87-4.78) and male gender (pOR 1.37, 95% CI 1.23-1.54) was associated with ICU mortality and male gender (pOR 1.8, 95% CI 1.25 – 2.59) with IMV mortality. Of symptoms and signs, only dyspnea was associated with ICU mortality (pOR 2.56, 95% CI 1.65 – 3.99). Of comorbidities, HTN was associated with both ICU and IMV mortality (pOR 2.02, 95% CI 1.37 – 2.98; pOR 1.5, 95% CI 1.06 - 2.12, respectively), whereas COPD (pOR 3.22, 95% CI 1.03 – 10.09), cardiovascular disease (CVD) (pOR 2.77, 95% CI 1.76 – 4.37), and DM (pOR 1.78, 95% CI 1.19 – 2.65) were associated with ICU mortality. Of clinical findings, we found obesity, white blood cell count (WBC), ALT, AST, creatinine, total bilirubin, D-dimer, prothrombin time (PT), C-reactive protein (CRP), creatine kinase, lactate, lactate dehydrogenase (LDH); and reduced PaO2/FiO2, albumin or lymphocyte count to be correlated with ICU mortality. Among complications, AKI (pOR 12.47, 95% CI 1.52 – 102.7) and ARDS (pOR 6.52, 95% CI 2.66 - 16.01) were associated with ICU mortality. Thrombocytopenia and lymphocytopenia were correlated with ICU mortality; and there were no reports with laboratory findings to allow IMV outcome analysis. Finally, IMV significantly correlated ICU mortality (pOR 16.46, 95% CI 4.37-61.96) based on 6 studies [10,11,18,20,31,33] with substantial heterogeneity (p-value <0.1, I^2^ > 60%) not explained by subgroup analysis. Eggers’ testing revealed no publication bias.

**Table 3.**
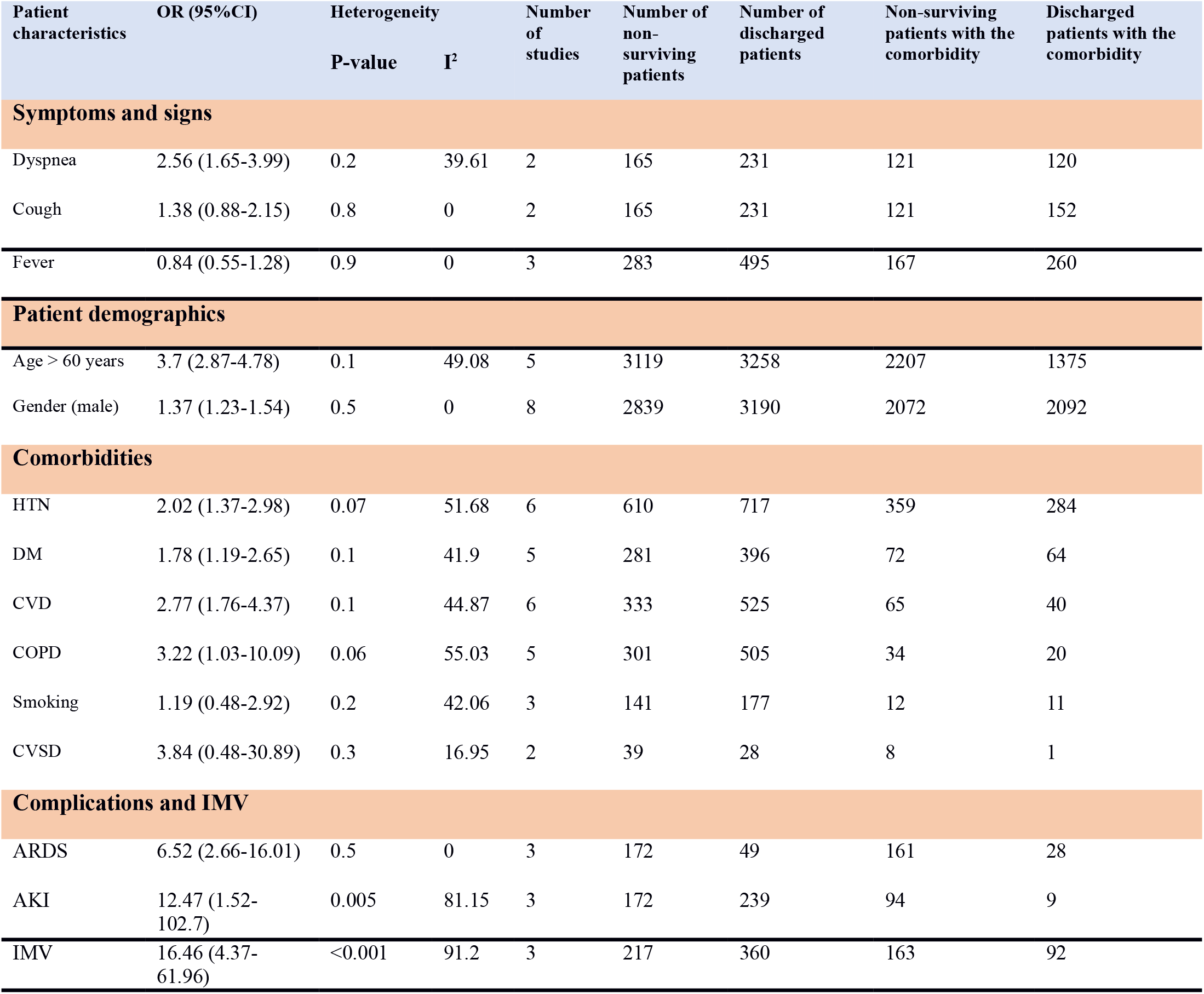
Meta-analysis of associations between COVID-19 ICU patient characteristics and ICU mortality. COVID-19 ICU patient characteristics including analysed demographics, presenting symptoms, comorbidities, complications and IMV and the corresponding number of non-surviving and discharged patients. Abbreviations: ARDS, adult respiratory distress syndrome; AKI, acute kidney injury; COPD, chronic obstructive pulmonary disease; CVD, cardiovascular disease; CVSD, cerebrovascular disease; DM, diabetes mellitus; HTN, hypertension, IMV, invasive mechanical ventilation.

**Table 4.** Meta-analysis findings of the association between laboratory findings and COVID-19 ICU mortality. Abbreviation: ALT, alanine aminotransferase; AST, aspartate aminotransferase; CRP, C-reactive protein; LDH, lactate dehydrogenase; WMD, weighted mean difference.

| Laboratory Finding | Number of studies | WMD (95% CI) | Meta-analysis P-value | Heterogeneity |  | Non-survived patients with data of the factor | Survived patients with data of the factor |
| --- | --- | --- | --- | --- | --- | --- | --- |
|  |  |  |  | P-value | I<sup>2</sup> |  |  |
| Oxygenation |  |  |  |  |  |  |  |
| Pao2/FiO2, mm/Hg | 2 | -33.763 (-46.936 - -20.591) | <0.001 | 0.8 | 0 | 84 | 149 |
| Cell Blood Count |  |  |  |  |  |  |  |
| Hemoglobin, g/dL | 2 | -0.380 (-1.348 – 0.587) | 0.44 | 0.02 | 71.60 | 143 | 278 |
| White blood cell count, x10<sup>9</sup>/L | 2 | 4.280 (3.675 – 4.885) | <0.001 | 0.5 | 0 | 244 | 469 |
| Lymphocyte count, x10<sup>9</sup> /L | 3 | -0.272 (-0.536 – -0.007) | 0.044 | <0.001 | 92.61 | 524 | 256 |
| Platelet count/μl | 3 | -16.073 (-57.853 – 25.707) | 0.45 | <0.001 | 90.69 | 276 | 489 |
| Coagulation index |  |  |  |  |  |  |  |
| D-dimer, ug/mL | 3 | 6.685 (1.328 – 12.042) | 0.014 | <0.001 | 95.49 | 242 | 467 |
| Prothrombin time, secs | 3 | 1.646 (1.362 – 1.930) | <0.001 | 0.2 | 33.326 | 228 | 328 |
| Inflammatory marker |  |  |  |  |  |  |  |
| CRP, mg/L | 3 | 86.217 (47.912 - 124.523) | <0.001 | <0.001 | 88.21 | 265 | 522 |

**Table 4. Meta-analysis findings of the association between laboratory findings and COVID-19**
| Biochemistry |  |  |  |  |  |  |  |
| --- | --- | --- | --- | --- | --- | --- | --- |
| Albumin, g/dL | 2 | -0.512 (-0.590 - -0.434) | <0.001 | 0.7 | 0 | 240 | 447 |
| ALT, U/L | 2 | 3.846 (-2.474 - 10.165) | 0.233 | 0.12 | 59.17 | 189 | 426 |
| AST, U/L | 2 | 22.996 (11.696 - 34.296) | <0.001 | 0.01 | 84.59 | 239 | 447 |
| Creatinine, mg/dL | 3 | 0.325 (-0.044 - 0.695) | 0.084 | <0.001 | 96.44 | 279 | 492 |
| Creatine kinase, U/L | 2 | 378.166 (-127.086 - 883.418) | 0.142 | <0.001 | 95.73 | 175 | 284 |
| Lactate, mmol/L | 2 | 0.289 (0.027 - 0.551) | 0.03 | 0.2 | 42.45 | 66 | 69 |
| LDH, U/L | 2 | 235.354 (174.931 - 295.778) | <0.001 | 0.01 | 84.63 | 200 | 399 |
| Total bilirubin, mmol/L | 2 | 5.317 (4.200 - 6.434) | <0.001 | 0.6 | 0 | 165 | 231 |

#### Regional differences in ICU and IMV outcomes

Subgroup analysis revealed the following regional ICU admissions: USA (35%), UK (17%) and China (14%); IMV rates: USA (85%), Italy (72%), France (69%), UK (66%), China (56%), Mexico (36%); ICU mortality rates: UK (33%), USA (29%), Italy (26%), China (24%), Spain (23%), France (15%), and Mexico (2%) and IMV mortality rates: China (59%) followed by UK (53%), USA (24%) and Mexico (4%).

## DISCUSSION

This is the most comprehensive systematic review and meta-analysis of COVID-19 ICU cases and outcomes to date to our knowledge. Many reviews explored COVID-19 prognosis, with some reportinggenerally on all [36] or hospitalized cases [37], or from a certain region [38], or focused on specific aspects such as mortality [39] or outcomes [40], with only one systematic review specifically addressing IMV mortality [41]. Here we attempt to provide a comprehensive assessment of ICU- admitted COVID-19 patient characteristics, regional discrepancies, as well as overall ICU and IMV specific outcomes and their associated factors.

Our pooled ICU best case mortality rate of 28.3% (95% CI 0.27-0.32) amongst COVID-19 patients is lower than earlier reports from Wuhan’s 78% [20] and Seattle’s 85% [8], but in line with the more recent report of 31% from Atlanta [10], as well as 25.7% and 41.6% reported in reviews by Quah et al. [39] and Armstrong et al. [40] respectively.

Our 43% (95% CI 0.29-0.58) pooled IMV mortality rate is also significantly lower than earlier reports of 86–97% from Wuhan [20.31] or 71-75% from Seattle [8.11]. The more recent Atlanta study revealed improved IMV mortality of 35.7% [10], which is more consistent with the 35% to 46% mortality respectively reported for patients intubated with H1N1 influenza pneumonia [42] and other causes of ARDS [43].

We found a pooled mean ICU LoS of 7.8 (7-8.5) days comparable to results from a review reporting a median LoS of 8 (5-13) days for patients from China and 7 (4-11) days from outside China [44]. As for IMV duration, we found a pooled mean duration of 10 days (95% CI 7-13), which is shorter than earlier reports from Wuhan and Seattle of 10-17 days [41].

Taken together, our results are consistent with the recent finding by Armstrong et alof improved COVID-19 ICU outcome over time, which was attributed to increased ICU experience, and evolving admission criteria, treatments, and demographics [40]. We additionally hypothesize that aimprovements in reported mortalities over time and regions may arise as healthcare systems become less overwhelmed, and as more recent studies account for more complete patient outcomes. Significant differences between reporting geographic regions could additionally be due to different ICU admission criteria and treatment protocols, diversity of ethnic genetic factors as well as mutational evolution of the virus itself [45], but more studies are necessary to explore these aspects.

Systematic reviews and analyses of select patient demographics, comorbidities, selective laboratory findings and complications of the disease have been published [46,47], although there is no systematic review addressing all these aspects in COVID-19 ICU or IMV to date. We found that elderly COVID-19 patients with comorbidities have higher ICU and IMV fatalities; but this is not unique to COVID-19 as many studies have noted that ICU patients generally consists of elderly patients who have greater mortality [48].

Interestingly, despite higher absolute prevalences of certain comorbidities and presenting features in COVID-19 ICU patients, the relative prevalence is the same as reported in less severe symptomatic or hospitalized cases. For example, a review of mostly Chinese studies similarly found hypertension to be the most common comorbidity in COVID-19 [49]; and another review identified fever (88.7%), cough (57.6%) and dyspnea (45.6%) to be similarly most common manifestations [50]. Our results also corroborated findings from meta-analyses where DM [51], CVD [52], abnormal liver function [53] have been found prevalent, associated and potentially prognostic in COVID-19.

For comorbidities, we found CVD, HTN and DM to be significantly associated with ICU mortality, which is consistent with findings by a review that identified these same comorbidities to be correlated with disease severity and ICU admissions [49]. Separately, Wang et al. also found HTM, DM, and CVD to correlate with COVID-19 severity and mortality [54].

Of clinical findings: obesity, WBC, AST, D-dimer PT, CRP, lactate, LDH, and total bilirubin as well as lower PaO2/FiO2, lymphocyte count and albumin to be significantly associated with ICU outcome (Table 4), which is again consistent with previous reports on associations of obesity, liver dysfunction, lymphocytopenia, coagulopathy, malnutrition, renal dysfunction, and cardiac injury with COVID-19 severity or mortality.

While most clinical correlates we found above are consistent and noted by other reports to date, three correlates with ICU mortality deserve discussion. One is AKI (pOR 12.47, 95% CI 1.52-102.7), for which there are divergent reports on prevalence and significance. Thus, while some large COVID-19 systematic reviews excluded AKI [27,37] and it is absent from some COVID-19 critical care management guidelines [55,56], its incidence in hospitalized COVID-19 has been reported to be as low as 0.5% [29] and some researchers even concluded that COVID-19 does not cause AKI [57]. However, we found a pooled AKI prevalence of 32% and a New York report cited 46% of hospitalized and 68% of ICU COVID-19 cases with AKI with 34% of those in ICU requiring replacement renal therapy (RRT), and with an OR of 20.9 (95% CI 11.7-37.3) for AKI vs non-AKI associated ICU mortality [58]. Along with our analysis, this raises the concern that AKI maybe seriously under appreciated in critically ill COVID-19 patients.

ARDS is another significantly associated complication (pOR 6.52, 95% CI 2.66 - 16.01) we found. Problematically, ARDS itself is a heterogeneous syndrome, with the six studies [8,9,22,31-33] we analyzed variously applying the Berlin criteria [59], WHO guidance [60], or no criteria at all for its diagnosis. Then there is a notion that COVID-19 ARDS may be a separate entity and that current criteria are inadequate in guiding treatment [61]. Indeed, ARDS per the Berlin criteria defines a one week onset limit whereas COVID-19 related ARDS has a median onset of 8-12 days [20]. Pathophysiologically, there are also differences between COVID or non-COVID ARDS and Gattinoni et al. have highlighted the non-uniformity of ARDS in COVID-19 based on elastance, ventilation/perfusion ratio, lung weight and recruitability [62]. Thus, further efforts are urgently needed in clarifying the pathophysiology and developing new stratifications of severity for COVID-19 specific ARDS to guide optimal ventilatory support for ICU COVID-19 patients.

Finally, the issue of defining ARDS in COVID-19 leads to applicability of IMV as its treatment. Early studies have reported ICU mortality rates as high as 86-97% for IMV in COVID-19 patients [18,20,31], but no study to date specifically reported on the relative odds for IMV v. non-IMV in terms of outcome. Our finding that IMV itself is significantly associated with ICU mortality (pOR 16.46, 95% CI 4.37-61.96) coupled with subgroup analyses showing regions with higher ICU mortalities coincide with regions with higher IMV rates (USA, UK, Italy) are notable because it questions the benefits of IMV as well as the wisdom of public policies emphasizing ventilator availability in coping with the pandemic. The concern for potentially harmful overuse of IMV has been raised by past research, since IMV itself carries risks that may adversely affect survival [63]. Additionally, inappropriate timing of intubation too early or too late, inadequately trained or overwhelmed staff, as well as improper ventilation settings and IMV associated pneumonia can all potentially enhance mortality; and this should all be investigated more fully in COVID-19 cases.

From prior studies on ARDS, we know that other ventilation strategies such as non-invasive ventilation (NIV) may improve mortality over IMV [64], and there are suggestions that non-rebreathing masks and prone positioning may improve outcome in hypoxemic COVID-19 patients [65]. The comparison of different ventilation strategies is beyond the scope of our review and there is yet no published study comparing ventilation strategies in COVID-19. However, our data supports a more conservative approach to initiating IMV, as well as the need for well designed trials to clarify the role of various ventilation strategies in COVID-19 associated ARDS.

Finally, our results highlight significant regional discrepancies in COVID-19 ICU and IMV rates and outcomes. As the pandemic evolves, regional differences of local health systems and their resources as well as genetic differences amongst populations and possible viral evolution over time can all account for such differences and deserves study. Regional differences also imply the need for flexible region-specific treatment protocols especially in resource-limited settings [65] instead of the aim to set uniform international protocols for treatment.

### Limitations

There are multiple limitations with our study. We have issues with incompleteness of data where not all outcomes are accounted for in many studies, as well as underlying heterogeneity of data with small number of studies and studies with small numbers of patients. Moreover, all our retrieved reports were retrospective cohorts, which limits our ability to infer causality; and some of these were not peer-reviewed. Furthermore, we could not adjust for confounders of potentially related variables in an analysis of survival vs. non-survival based on these studies. Finally, regional differences in health systems and their different treatment protocols may also confound results.

### Conclusions

This is the largest and most comprehensive review and meta-analysis of ICU and IMV outcomes in COVID-19 ICU patients. Our findings parallel earlier reports on prevalences of associated comorbidities and clinical findings in COVID-19 patients implying largely the same set of factors is associated with severity and outcome regardless of stage of disease. However, we highlight the significant association of AKI and ARDS associated IMV in ICU outcomes, which deserve further research to refine diagnostic criteria and enable the development of optimally tailored treatment strategies, as well as better planning and allocation of critical care resources. Finally significant regional discrepancies in outcomes implies a need for further studies to allow a broader perspective of factors associated with ICU and IMV risks and outcomes under different healthcare systems in different populations.

## Data Availability

All data referred to in the manuscript is available from the corresponding author.

**Abbreviations**
AKI: acute kidney injury;
ALT: alanine aminotransferase;
AST: aspartate aminotransferase;
ARDS: acute respiratory distress syndrome;
CI: confidence interval;
COPD: chronic obstructive pulmonary disease;
COVID-19: coronavirus disease of 2019;
CVD: cardiovascular disease;
CRP: C-reactive protein;
DM: diabetes mellitus;
HTN: hypertension;
ICNARC: Intensive Care National Audit and Research Center;
ICU: intensive care unit;
IMV: invasive mechanical ventilation;
LDH: lactate dehydrogenase;
LoS: length of stay;
pER: pooled event rate;
pOR: pooled odds ratio;
PT: prothrombin time;
RRT: renal replacement therapy;
SARS-CoV-2: severe acute respiratory syndrome coronavirus 2;
WBC: white blood cell count;
WHO: World Health Organization.

## Supporting Information

**S1 Table. Prisma Checklist**

**S1 File. PROSPERO Registration**

